# Preprint: Towards Smart Glasses for Facial Expression Recognition Using OMG and Machine Learning

**DOI:** 10.1101/2023.04.14.23288552

**Authors:** Ivana Kiprijanovska, Simon Stankoski, M. John Broulidakis, James Archer, Mohsen Fatoorechi, Martin Gjoreski, Charles Nduka, Hristijan Gjoreski

**Affiliations:** Emteq Ltd., Brighton BN1 9SB, UK; Faculty of Informatics, Università della Svizzera Italiana, 6900 Lugano, Switzerland; Faculty of Electrical Engineering and Information Technologies, Ss. Cyril and Methodius University in Skopje, 1000 Skopje, North Macedonia

**Author notes:** Correspondence (I.K). (S.S); (M.J.B); (J.A); (M.F.); (C.N). (M.G). (H.G.).

## Abstract

This study aimed to evaluate the use of novel optomyography (OMG) based smart glasses, OCOsense™, for the monitoring and recognition of facial expressions. Experiments were conducted on data gathered from 27 young adult participants, who performed facial expressions varying in intensity, duration, and head movement. The facial expressions included smiling, frowning, raising the eyebrows, and squeezing the eyes. The statistical analysis demonstrated that: (i) OCO™ sensors based on the principles of OMG can capture distinct variations in cheek and brow movements with a high degree of accuracy and specificity; (ii) Head movement does not have a significant impact on how well these facial expressions are detected. The collected data were also used to train a machine learning model to recognise the four facial expressions and when the face enters a neutral state. We evaluated this model in conditions intended to simulate real-world use, including variations in expression intensity, head movement and glasses position relative to the face. The model demonstrated an overall accuracy of 93% (0.90 f1-score) – evaluated using a leave-one-subject-out cross-validation technique.

## Introduction

Affective computing and remote emotion monitoring are scientific fields that can have a strong impact on digital medicine, especially in the realm of digital solutions for mental-health management [1]. The applicability of such digital solutions extends beyond clinical studies, given that the World Health Organization (WHO) considers mental health as more than simply the absence of mental disorders. It exists on a complex continuum, experienced differently from one person to another [2]. Thus, it would be extremely beneficial if people could monitor clinically significant parameters known to be related to mental health (e.g., facial expressions and emotions) with the same ease as they can track heart rate and daily step count. Qualitative mental health management would also have a positive effect on cognitive aging, as poor mental health accelerates age-related cognitive decline [3]. Cognitive aging is a worldwide emergency in and of itself. In 2020, more than 20.6% of the EU population was aged over 64, and this percentage will increase by at least 3% every ten years [4]. In vulnerable populations with diagnosed medical problems, mental health disorders (such as depression) double the risk for cardiac mortality in people with and without cardiac disease. This relation has been confirmed for major depression, as well as in volunteers with elevated depressive traits, even when they fail to meet a formal diagnosis [5]. Another study linked depression symptoms and ischemic heart disease (coronary heart, heart attack, and angina) [6].

The face is one of the most expressive parts of our body [7] and has an important role in communicating emotional and mental states, as well as behavioural intentions. The relationship between facial expressions and emotions has been studied since the late 19^th^ century [8]. In the late 20^th^ century Ekman’s ground-breaking studies first hypothesized that emotions and facial expressions are universally recognizable [9]. In recent years, this direct mapping between a facial expression and a given emotion has been significantly challenged. Meta-analyses of autonomic physiology, behaviour, and even brain imaging, all report little evidence for consistent and specific facial expression derived ‘fingerprints’ for different categories of emotion, like anger, sadness, and fear [10, 11]. Instead, whilst emotions and affective states are related to facial expressions, the relationship is likely both personalized and context-based [12]. Thus, one should be careful when analyzing emotional states through the lenses of facial expression recognition systems that assume a direct correspondence between expressions and emotions.

Given the relationship between facial expressions and mental states, a variety of remote sensing approaches have been proposed in the past [13, 14]. Regarding the recent studies related to head-worn sensors for facial tracking, Sato et al. [15] and Gjoreski et al. [16] investigated wearable solutions based on electromyography (EMG) sensors that are positioned over specific facial muscles: the frontalis (left and right side of the forehead; the orbicularis (left and right side of the eyes); the zygomaticus (left and right side of the cheeks); and the corrugator muscle (between the eyebrows). Yan et al. [17] explored expression recognition with a glasses camera facing the wearer, and Matthies et al. [18] explored glasses-based capacitive sensing for tracking facial expressions and gestures. Regarding the commercially available smart glasses, the OCO™ sensors are always oriented towards the user/wearer, thus avoiding one of the biggest privacy issues that smart glasses have. For example, Google Glass – and similar solutions from Sony (SmartEyeGlass) [19] and Vuzix (M300) [20] – introduce privacy concerns as these devices could continuously record video using a front-mounted camera, compromising bystanders’ privacy [21]. Regarding other face-tracking tools, camera-based systems are frequently used to track facial expressions and infer emotional states in the wild. However, their effectiveness is limited due to restrictions on sensor location, lighting, intrusiveness, and movement.

This study investigates the feasibility of using novel optomyography (OMG) based glasses for recognizing four facial expressions: smile, frown, eyebrow raise and squeezed eyes. Activation of the muscles involved in creating these expressions is used to determine emotional response, specifically valence [22] and pain [23]. The OCOsense™ system consists of multi-sensor wearable glasses (Figure 1). The OCO™ sensors are non-contact optical sensors that can read facial skin movement in three dimensions, providing a higher resolution signal (±4.7μm X&Y-axis, ±4.0μm Z-axis) than the average camera-based solution and has advantages over EMG-based systems. Although EMG is perhaps the most sensitive way to objectively track facial movement, the electrodes also require firm and constant contact with the skin in order to achieve an acceptable signal-to-noise ratio, which is not practical in a glasses format. Instead, the OCO™ sensors are optically based, removing the requirement for direct skin contact and are able to function accurately from 4 mm to 30 mm away from the skin.

**Figure 1.**
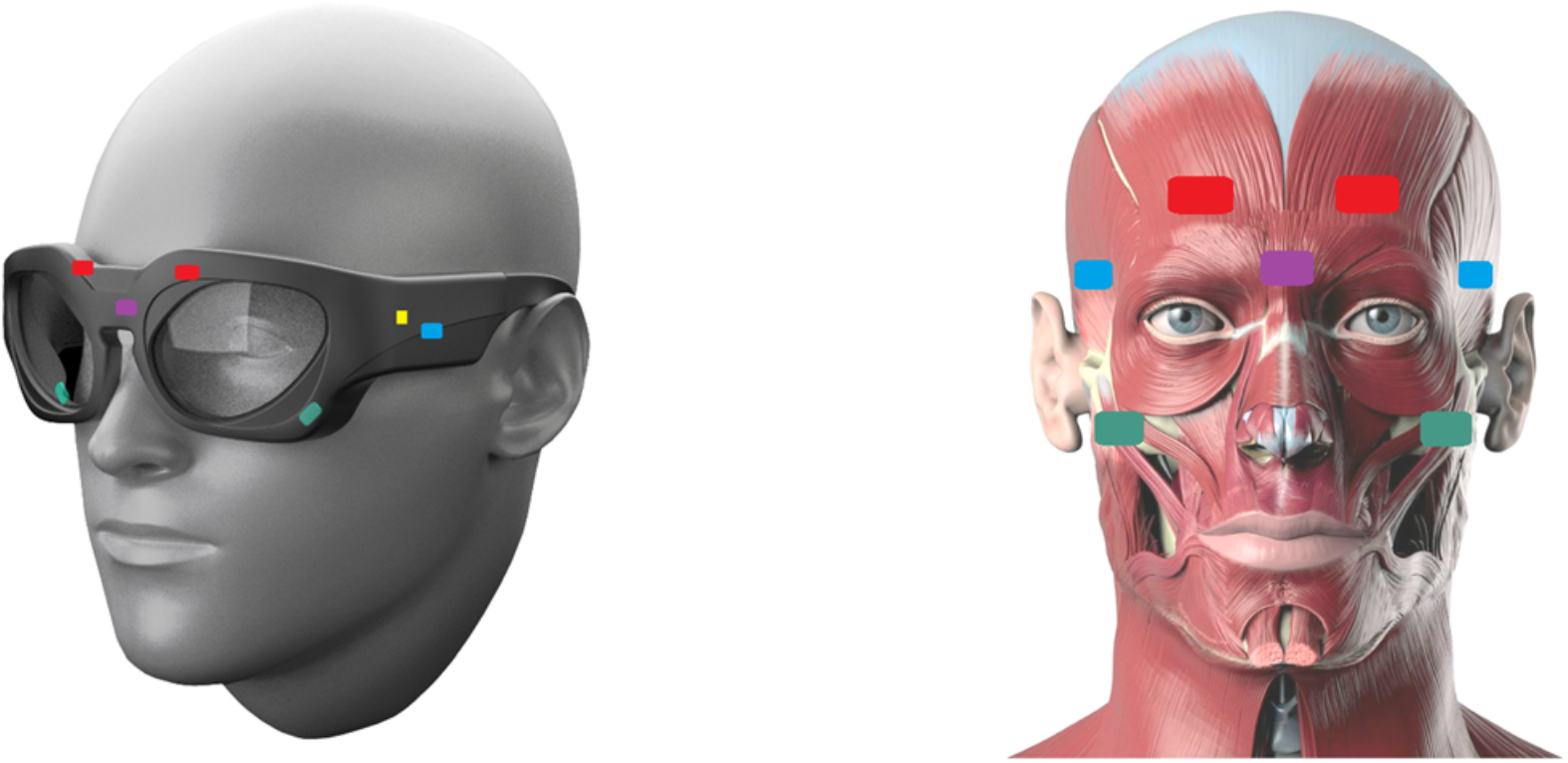
OCOsense™ glasses and corresponding sensor placement. The coloured rectangles represent the OCO™ sensors within the frame, over major facial muscles: frontalis (red), zygomaticus (green), and corrugator (purple). The blue OCO™ sensors are positioned over the temples. Also, there is a 9-axis IMU and altimeter (yellow). Source of the right image [24].

More specifically, in this study we present analysis about:

- The relationship between the OCO™ sensor data and facial expressions.
- The influence of head movement on OCO™ sensor data.
- The influence of glasses position on OCO™ sensor data.
- The usage of machine learning for facial expression recognition from OCO™ sensor data.

The experimental results demonstrated that the OCO™ sensors can capture distinct variations in the cheek and brow movements for each of the facial expressions studied. Specifically, cheek movements were primarily observed during the smile and squeezed eyes expression, in contrast to the frown and eyebrow raise expression. Conversely, brow movements were primarily detected during the eyebrow raise and frown expression, compared to the squeezed eyes and smile expression. The study also found head movement does not have a significant impact on the skin movement measurements obtained by the OCO™ sensors during different facial expressions. However, the position of the glasses on the face does, particularly when monitoring brow movement during a frown, eyebrow raise or squeezed eyes expression. The measurements obtained from the OCO™ sensors were also used to train a machine learning model to recognize smile, frown, eyebrow raise, and squeezed eyes expression, and when the face enters a neutral state. The model evaluated different conditions expected to mimic real-world use, including variations in expression intensity, head movement and glasses position, demonstrating an overall accuracy of 93% (0.90 f1-score). These findings highlight the ability of the OCO™ sensors to accurately capture facial movements associated with different facial expressions in various conditions, providing evidence for their potential use as a reliable tool for monitoring and detecting changes in facial expressions.

## Results

### Relationship between facial expressions and OCO™ sensor data

#### Low-intensity expressions

To assess the ability of the OCO™ sensors to detect movement of facial muscles during different facial expressions, we conducted statistical analyses, which involved comparing the measurements of the brow and cheek sensors during different voluntary facial expressions. To perform statistical tests, from the brow and cheek OCO™ sensors (right and left cheek, right and left eyebrow) measuring the skin movement along the X-, Y-, and Z-axis, we calculated the mean cheek and brow movements for each expression type, for each subject. For the statistical analysis on the low-intensity expressions, we used the data where the participants were performing both short (1 second) and long (3 seconds) low-intensity expressions, and the mean movement for a specific expression type was calculated over all performed expressions of that type by each participant. This procedure led to n=27 (number of participants in the dataset) tuples of four values, where each value represented the mean cheek/brow movement for each expression type (eyebrow raise, frown, smile, and squeezed eyes). To test if there is a statistically significant difference in the mean cheek and brow movements detected by the glasses for all pairs of performed expressions – eyebrow raise vs. frown, eyebrow raise vs. smile, eyebrow raise vs. squeezed eyes, frown vs. smile, frown vs. squeezed eyes, and smile vs. squeezed eyes – we used the Wilcoxon signed-rank (paired) test with an with Bonferroni correction (alpha = 0.05). Figure 2 shows the mean cheek (left plot) and brow (right plot) movements during different low-intensity expressions, presented on the X-axis (from left to right: eyebrow raise, frown, smile, squeezed eyes), and the results from the statistical test.

**Figure 2.**
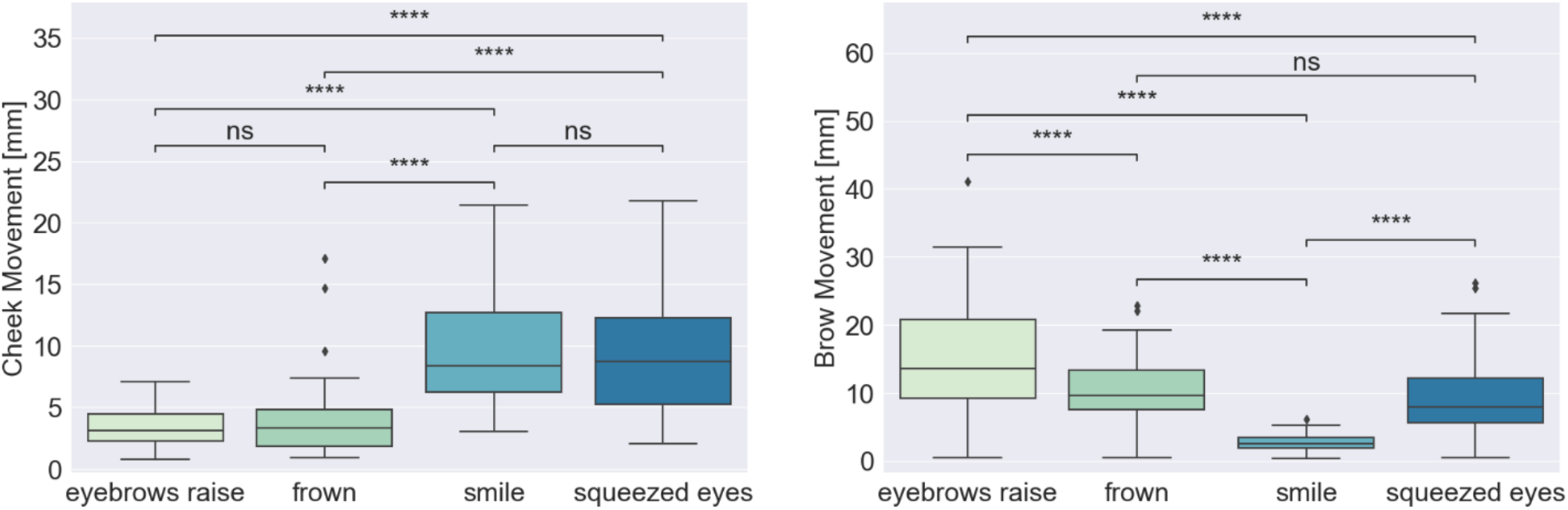
Wilcoxon signed-rank (paired) test with Bonferroni correction for comparing mean cheek and brow movements during low-intensity expressions pairs: eyebrow raise vs. frown, eyebrow raise vs. smile, eyebrow raise vs. squeezed eyes, frown vs. smile, frown vs. squeezed eyes, and smile vs. squeezed eyes. Statistical significance annotations: * if p ∈ [.05, 10^−2^); ** if p ∈ [10^−2^, 10^−3^); *** if p ∈ [10^−3^, 10^−4^); and **** if p ≥ 10^−4^.

For the cheek OCO™ sensors, we can observe more movement during the smile and squeezed eyes expression (median values 5.84 mm and 5.91 mm, respectively) compared to during eyebrow raise and frown expression (median values 2.25 mm and 2.48 mm, respectively). The results from the statistical test indicate that the mean movement of the cheek is significantly different between eyebrow raise and smile expression (p-value: 2.505×10^−9^), eyebrow raise and squeezed eyes expression (p-value: 3.983×10^−9^), frown and smile expression (p-value: 4.415×10^−7^), and frown and squeezed eyes expression (p-value: 4.909×10^−8^). The difference in cheek mean movement between eyebrow raise and frown expression (p-value: 1×10^0^) and between smile and squeezed eyes expression (p-value: 1×10^0^) is not statistically significant.

For the brow OCO™ sensors, we can observe the most movement during the eyebrow raise expression (median value: 10.14 mm), followed by the frown and squeezed eyes expression (median values 7.46 mm and 5.70 mm, respectively), while the lowest movement is observed during the smile expression (median value: 1.84 mm). The results from the statistical test indicate that the movement of the brow is significantly different between all pairs of expressions, except the frown and squeezed eyes expression (p-value: 1×10^0^). The p-values for the rest of the expression pairs are lower than 10^−4^.

#### High-intensity expressions

For the statistical analysis on the high-intensity expressions, we used data where the participants were performing both short (1 second) and long (3 seconds) high-intensity expressions, and the mean movement for a specific expression type was calculated over all performed expressions of that type by each participant. This procedure led to n=27 (number of participants in the dataset) tuples of four values, where each value represented the mean cheek/brow movement for each expression type (eyebrow raise, frown, smile, and squeezed eyes). To test if there is a statistically significant difference in the mean cheek and brow movements detected by the glasses for all pairs of performed expressions – eyebrow raise vs. frown, eyebrow raise vs. smile, eyebrow raise vs. squeezed eyes, frown vs. smile, frown vs. squeezed eyes, and smile vs. squeezed eyes – we used the Wilcoxon signed-rank (paired) test with Bonferroni correction (alpha = 0.05). Figure 3 shows the mean cheek (left plot) and brow (right plot) movements during different high-intensity expressions, presented on the X-axis (from left to right: eyebrow raise, frown, smile, squeezed eyes), and the results from the statistical test.

**Figure 3.**
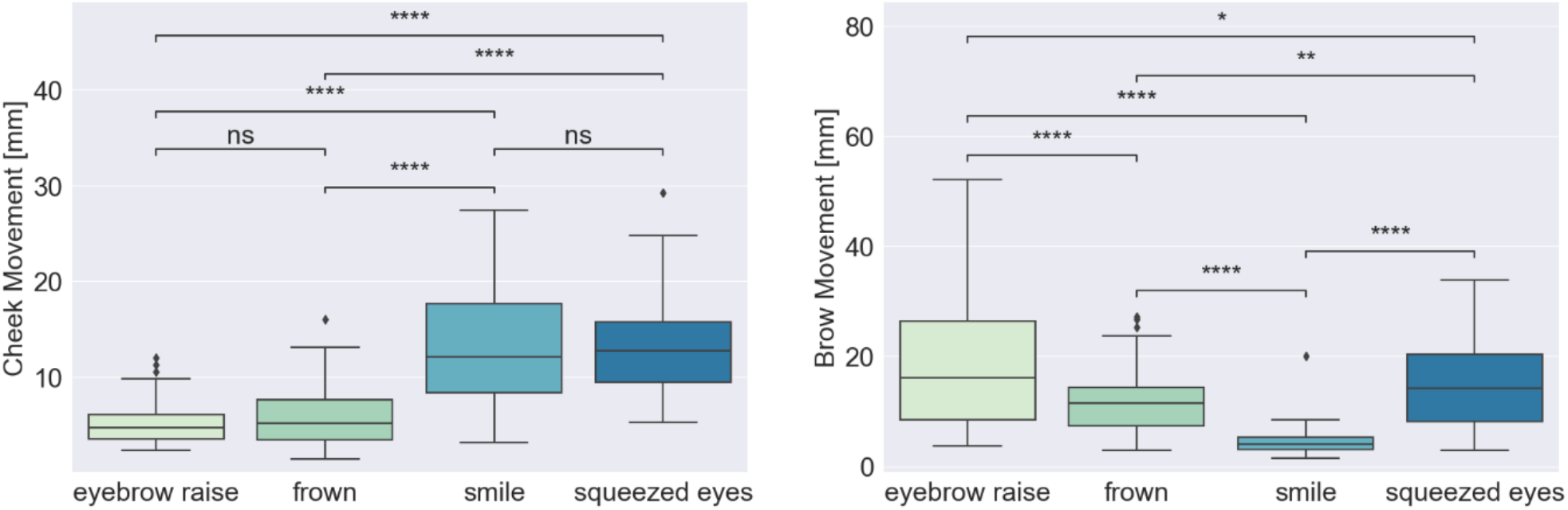
Wilcoxon signed-rank (paired) test with Bonferroni correction for comparing mean cheek and brow movements during high-intensity expressions pairs: eyebrow raise vs. frown, eyebrow raise vs. smile, eyebrow raise vs. squeezed eyes, frown vs. smile, frown vs. squeezed eyes, and smile vs. squeezed eyes. Statistical significance annotations: * if p ∈ [.05, 10^−2^); ** if p ∈ [10^−2^, 10^−3^); *** if p ∈ [10^−3^, 10^−4^); and **** if p ≥ 10^−4^.

For the cheek OCO™ sensors, we again observe more movement during the smile and squeezed eyes expression (median values 12.14 mm and 12.70 mm, respectively) compared to during the eyebrow raise and frown expression (median values 4.70 mm and 5.17 mm, respectively). The results from the statistical tests indicate that the cheek movement is significantly different between eyebrow raise and smile expression (p-value: 1.640×10^−8^), eyebrow raise and squeezed eyes expression (p-value: 2.102×10^−9^), frown and smile expression (p-value: 9.820×10^−7^), and frown and squeezed eyes expression (p-value: 2.553×10^−8^). The difference in cheek movement between eyebrow raise and frown expression (p-value: 9.011×10^−1^) and between smile and squeezed eyes expression (p-value: 1×10^0^) is not statistically significant.

For the brow OCO™ sensors, we can observe the highest movement intensity during the eyebrow raise expression (median value: 16.16 mm), followed by the squeezed eyes and frown expression (median values 14.14 mm and 11.51 mm, respectively), while the lowest movement intensity is observed during the smile expression (median value: 4.00 mm). The results from the statistical test indicate that the movement of the brow is significantly different between all pairs of expressions. The lowest difference is observed between the eyebrow raise and squeezed eyes expression (p-value: 2.473×10^−2^), followed by the frown and squeezed eyes expression (p-value: 1.132×10^−3^). The p-values for the rest of the expression pairs are lower than 10^−4^.

### Influence of head movement on OCO™ sensor data

To investigate the influence of head movement on the OCO™ sensor data, we performed statistical tests on the mean cheek and brow movements differences between expressions performed while the participants held their head still, and expressions performed while the participants simultaneously moved their head in a specific direction (to the right, to the left, upwards, or downwards). For these comparisons, we used only high-intensity, long-duration (3 seconds) expressions data, and the mean movement for a specific expression type was calculated over all performed expressions of that type by each participant. This procedure led to n=27 (number of participants in the dataset) tuples of two values for each expression type, for both the cheek and the brow movement. One of the tuples values is representing the mean value for the expressions where no head movement was included, and the other one for the expressions performed with head movement included. To test if there is a statistically significant difference in the mean cheek and brow movements detected by the glasses for all expressions in the movement and no-movement condition we used the Wilcoxon signed-rank (paired) test with Bonferroni correction (alpha = 0.05). Figure 4 shows the mean cheek (left plot) and brow (right plot) movements during different high-intensity expressions performed with/without head movement, presented on the X-axis (from left to right: eyebrow raise, frown, smile, squeezed eyes), and the results from the statistical test.

**Figure 4.**
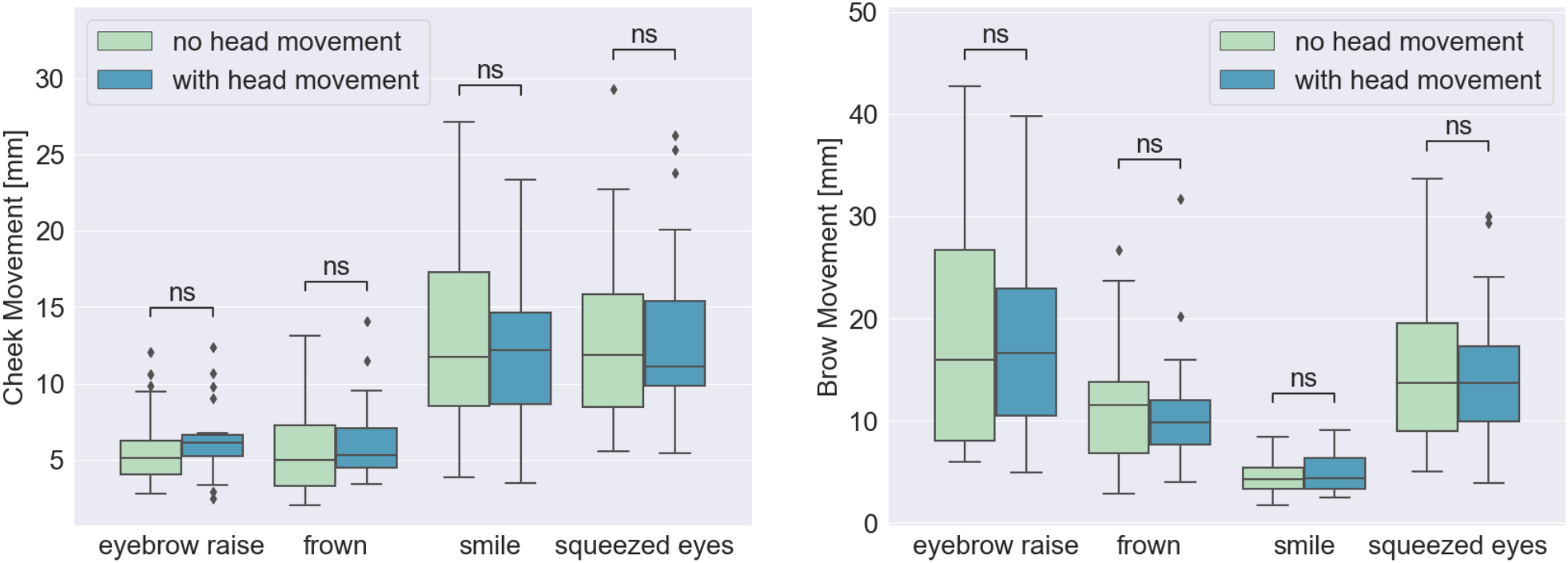
Wilcoxon signed-rank (paired) test with Bonferroni correction for cheek and brow movements between expressions (eyebrow raise, frown, smile, and squeezed eyes) performed with and without head movement. Statistical significance annotations: * if p ∈ [.05, 10^−2^); ** if p ∈ [10^−2^, 10^−3^); *** if p ∈ [10^−3^, 10^−4^); and **** if p ≥ 10^−4^.

For the cheek movements, the statistical test showed p-values higher than the significance level of 0.05 for all expression types (p-value=8.690×10^−1^ for the eyebrow raise expression, p-value=3.973×10^−1^ for the frown expression, p-value=4.421×10^−1^ for the smile expression, and p-value=1×10^0^ for the squeezed eyes expression) indicating that there is no statistically significant difference between the two analysed conditions. The same was confirmed with the statistical tests applied on the brow sensor measurements, which also showed p-values higher than the significance level of 0.05 for all expression types (p-value=1×10^0^ for the eyebrow raise, frown, smile, and squeezed eyes expression).

### Influence of glasses position on OCO™ sensor data

To evaluate the influence of the positioning of the glasses on the OCO™ sensor data during different expressions, we performed statistical tests on the mean cheek and brow movements differences between expressions of high-intensity and short (1 second) and long (3 seconds) duration performed while the participants were wearing the glasses in three different positions: *low* (the lowest point on the nasal bridge where the glasses are staying in place), *medium* (the medium point on the nasal bridge), and *high* (the highest point on the nasal bridge). The mean movement for a specific expression type was calculated over all performed expressions of that type by each participant. This procedure led to n=27 (number of participants in the dataset) tuples of three values for each expression type, for both the cheek and the brow movement. One of the tuples values is representing the mean value for the expression performed for the low position, the other is representing the mean value for the expression performed for the medium position, and the third one is related to the high position. To test if there is a statistically significant difference in the mean cheek and brow movements detected by the glasses for all expressions performed while the participants were wearing the glasses in three distinct positions, we used the Wilcoxon signed-rank (paired) test with Bonferroni correction (alpha = 0.05). Figure 5 shows the mean cheek (left plot) and brow (right plot) movements during different expressions performed while the participants were wearing the glasses in the three distinct positions, and the results from the statistical test.

**Figure 5.**
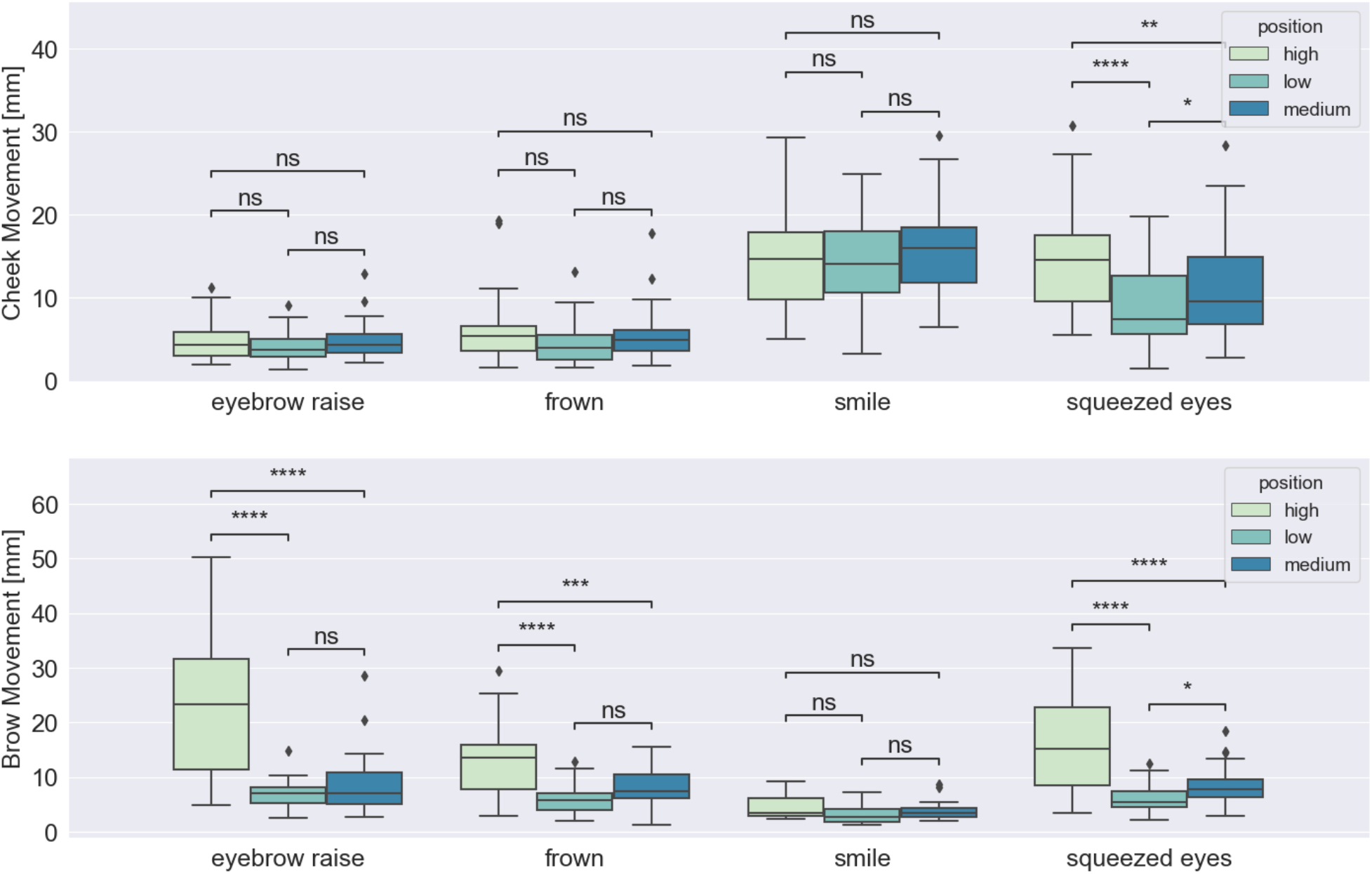
Wilcoxon signed-rank (paired) test with Bonferroni correction for cheek and brow movements between expressions (eyebrow raise, frown, smile, and squeezed eyes) performed while wearing the glasses in different positions on the nose bridge (low, medium, and high). Statistical significance annotations: * if p ∈ [.05, 10^−2^); ** if p ∈ [10^−2^, 10^−3^); *** if p ∈ [10^−3^, 10^−4^); and **** if p ≥ 10^−4^.

The statistical test results showed no statistically significant difference in the cheek movement between all three positions of the glasses during the eyebrow raise and frown expression, which are characterized with no notable cheek activations. The results also showed that there is no significant difference in the cheek movements detected by the sensors between all three positions of the glasses during the smile expression. For the squeezed eyes expression, the measured cheek movements were significantly different in the three cases: high vs. medium position (p-value=8.013×10^−3^), high vs. low position (p-value=3.701×10^−5^), and medium vs. low position (p-value=2.024×10^−2^).

On the other hand, for the brow movements, the statistical test results showed that the position of the glasses has no influence on the measurements only during the smile expression. For the frown expression, there was a significant difference between the high and low position (p-value=7.993×10^−5^) and a significant difference between the high and medium position (p-value=7.598×10^−4^). Comparable results were obtained for the eyebrow raise expression. Namely, when the glasses were positioned highest on the nose bridge, the detected brow movements were significantly higher compared to when the glasses were positioned low or medium on the nose bridge (p-value=8.941×10^−7^, and p-value=1.788×10^−7^, respectively). Lastly, for the squeezed eyes expression, there was a significant difference between all tested pairs of glasses positioning: high vs. low position (p-value=7.689×10^−6^), high vs. medium position (p-value=1.788×10^−6^), and medium vs. low position (p-value=2.454×10^−2^).

### Machine learning for facial expression recognition from OCO™ sensors data

The same dataset from the statistical analysis, was also used to train a machine learning model for recognizing facial expressions and when the face enters a neutral state. The dataset contains four distinct facial expressions: smile, frown, eyebrow raise, and squeezed eyes, with varying degrees of intensity and duration. Additionally, a subset of the expressions was recorded while the participants were simultaneously moving their head in a specific direction. All participants performed the expressions while wearing the glasses in one of three different positions on the nose: low, medium, and high. After the segmentation and feature extraction, the dataset contained 361,226 instances and 162 features, which were fed to a Random Forest model. Figure 6 presents the distribution of the labels of the machine learning instances. More details about the machine learning pipeline are presented in the section *OCOsense™ Expression Recognition Machine Learning Pipeline*. The focus in these experiments was on the data rather than the machine learning pipeline. Because of that, the pipeline was simplistic in order to avoid high evaluation scores because of advanced data processing and machine learning modelling.

**Figure 6.**
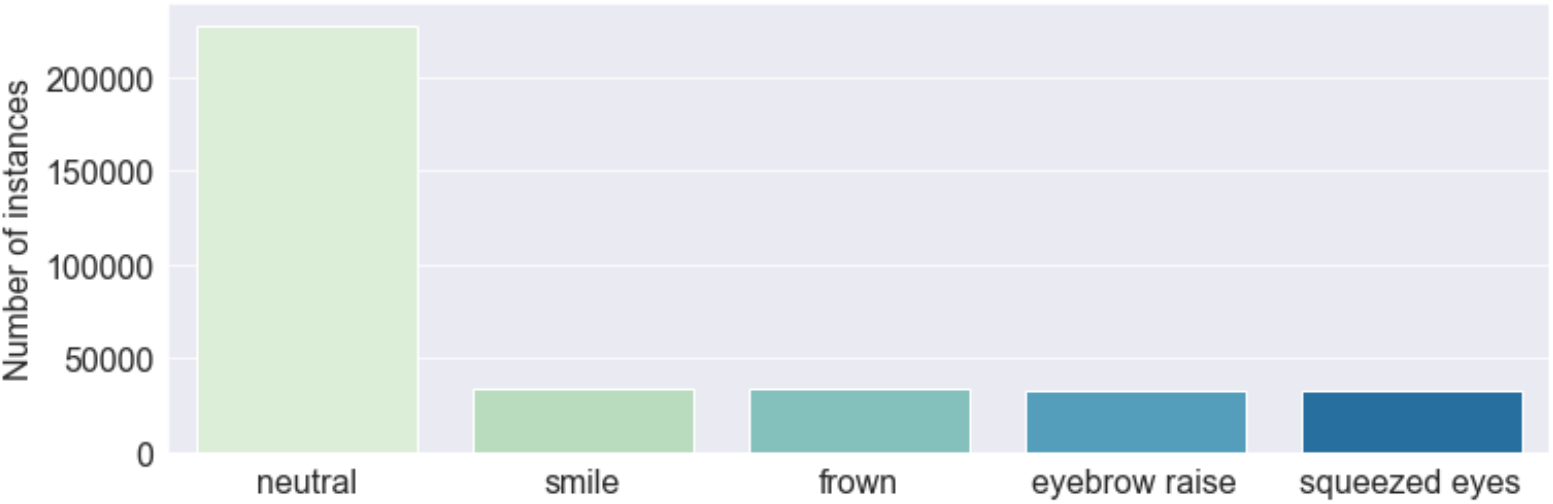
Distribution of expression labels in the dataset.

The machine learning model yielded an overall accuracy of 93% (0.90 f1-score), evaluated on the data collected from n=27 participants using a leave-one-subject-out cross-validation technique. The classification matrix is illustrated in Figure 7, and the classification report is shown in Table 1. The model achieved high evaluation scores for recognizing smile, eyebrow raise, and squeezed eyes expression, with recall scores of 0.9 for each of these expressions. The lowest recall score is observed for the frown expression, which is mistaken for the expression of squeezed eyes in 10% of the cases. This misclassification can be attributed to the similarities in the movement of the eyebrow during these expressions, which are both characterized by the activation of the frontalis and corrugator supercilii muscle, leading to the lowering of the inner portion of the eyebrow and the bringing together of the eyebrows. Another notable aspect of the model’s performance is that, aside from the misclassification of the frown expression, the model demonstrates minimal instances of confusion between other expressions. This suggests that the differences in the cheek and brow movements during different expressions are easily detectable by the sensors in the glasses.

**Table 1.**
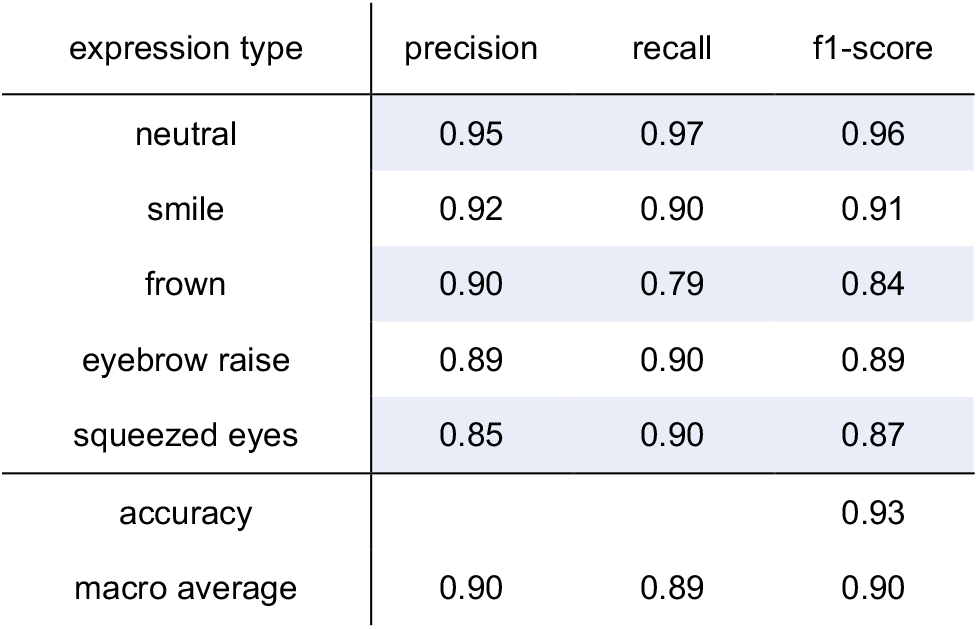
Classification report for the ML facial expression recognition model evaluated on the whole dataset.

**Figure 7.**
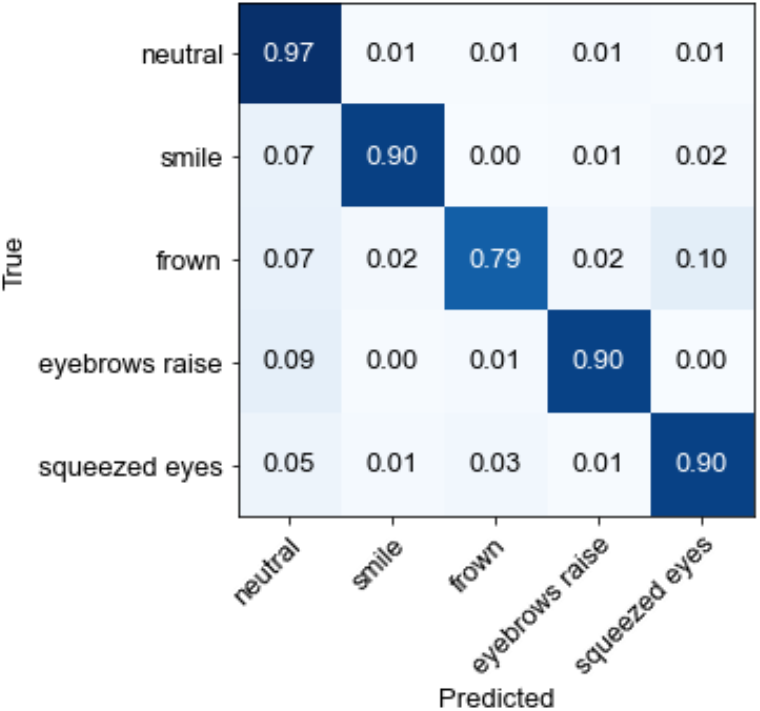
Confusion matrix for the ML facial expression recognition model evaluated on the whole dataset.

#### Low-intensity expressions vs. high-intensity expressions

To investigate the effect of expression intensity on the model’s performance in recognizing facial expressions, separate analyses were conducted on low-intensity and high-intensity expression data. The classification matrices for the low-intensity and high-intensity expressions are illustrated in Figure 8 and Figure 9, respectively. The classification reports are also presented in Table 2 and Table 3. From the analyses, it can be observed that the model demonstrates only slightly better performance for the recognition of high-intensity expressions, achieving an accuracy of 95% (0.92 f1-score), compared to the low-intensity expressions, where it achieves 92% accuracy (0.88 f1-score). While the rate of misclassification between different expressions is consistent across both data sets, a lower rate of instances of expressions being classified as neutral was observed for high-intensity expressions. Overall, the results indicate that the glasses sensors can capture even minimal muscle activations that are related to different facial expressions, which are sufficient for the model to recognize the performed expression.

**Table 2.**
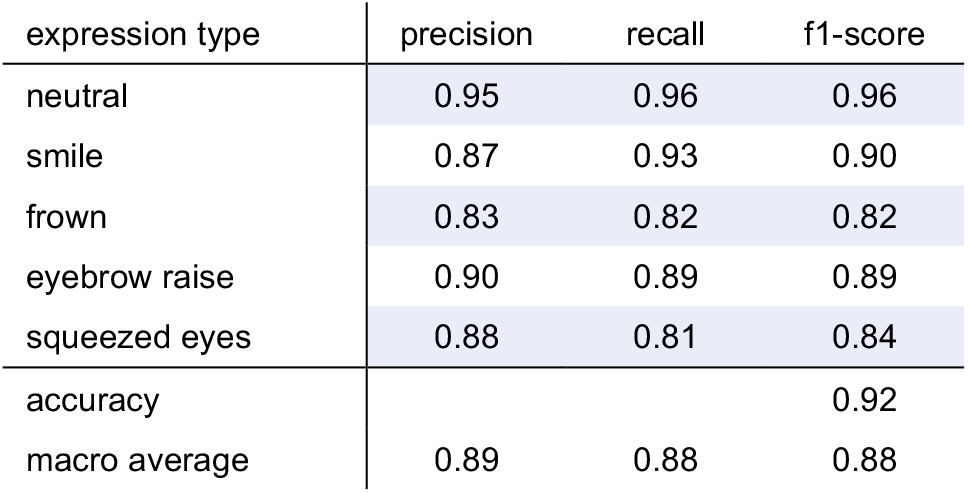
Classification report for the ML facial expression recognition model evaluated on the low-intensity facial expressions.

**Table 3.**
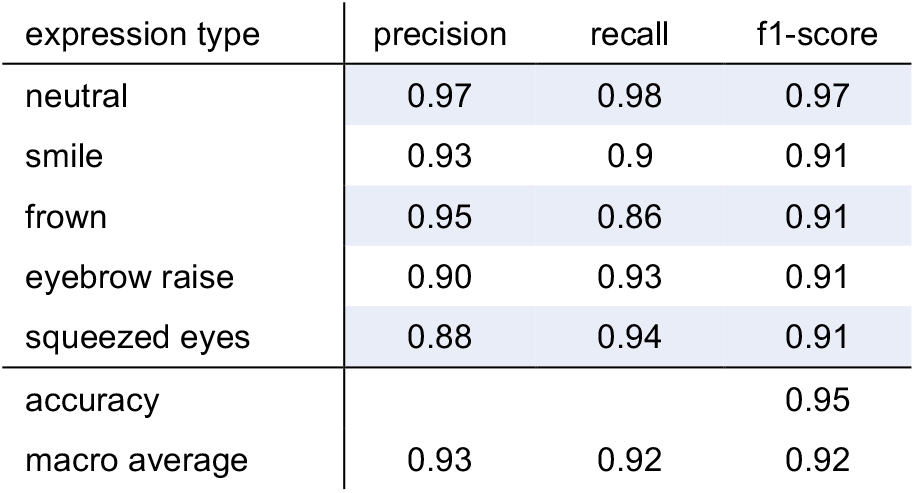
Classification report for the ML facial expression recognition model evaluated on the high-intensity facial expressions.

**Figure 8.**
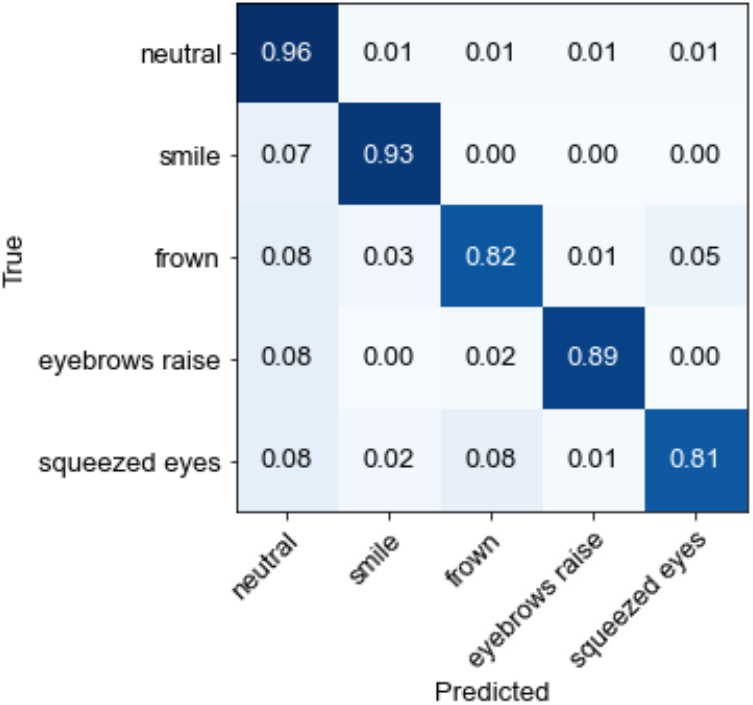
Confusion matrix for the ML facial expression recognition model evaluated on the low-intensity facial expressions.

**Figure 9.**
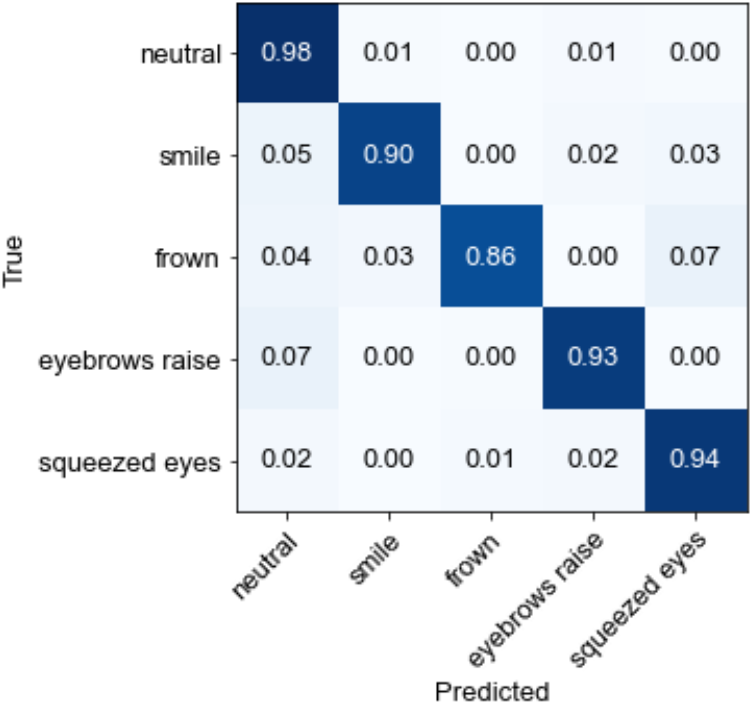
Confusion matrix for the ML facial expression recognition model evaluated on the high-intensity facial expressions.

#### No-movement vs. movement expressions

To investigate the influence of head movement on the model’s performance in recognizing facial expressions, separate analyses were conducted on expressions performed while the participants had their head still, and expressions performed while the participants simultaneously moved their head in a specific direction (to the right, to the left, upwards, or downwards). These comparisons include only high-intensity, long-duration expressions data. The classification matrices for both cases are illustrated in Figure 10 and Figure 11. The classification reports are also presented in Table 4 and Table 5. The analyses show that the model demonstrates only slightly better performance for the recognition of facial expressions performed while participants had their head still, achieving 95% accuracy (f1-score 0.93), compared to the expressions performed while participants moved their head, where it achieves 92% accuracy (0.90 f1-score). Again, the rate of misclassification between different expressions is low and consistent across both data sets, exept in the frown expression case. Namely, the model correctly identifies 89% of the frown expression instances in the no-movement scenario, while misclassifing only 4% as squeezed eyes expression, compared to 80% of correctly identified frown expression instances in the movement scenario, and 8% instances misclassified as squeezed eyes expression. Overall, the results indicate that the OCO™ sensors incorporated in the glasses are not substantially influenced by head movement and are able to capture facial movements related to different facial expressions even in motion scenarios. This eventually results in a robust machine learning model for facial expression recognition that has a high accuracy and consistency, regardless of the inclusion of head movement.

**Table 4.**
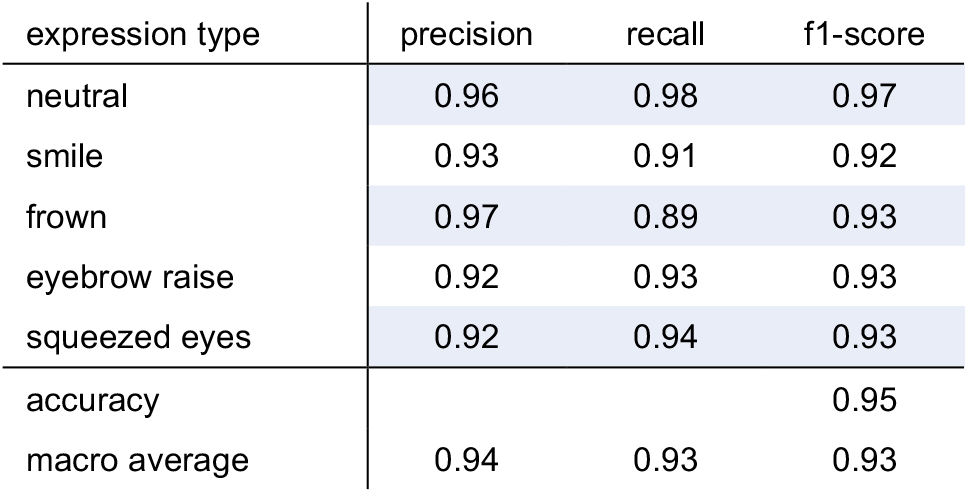
Classification report for the ML facial expression recognition model evaluated on the facial expressions performed in a still scenario.

**Table 5.**
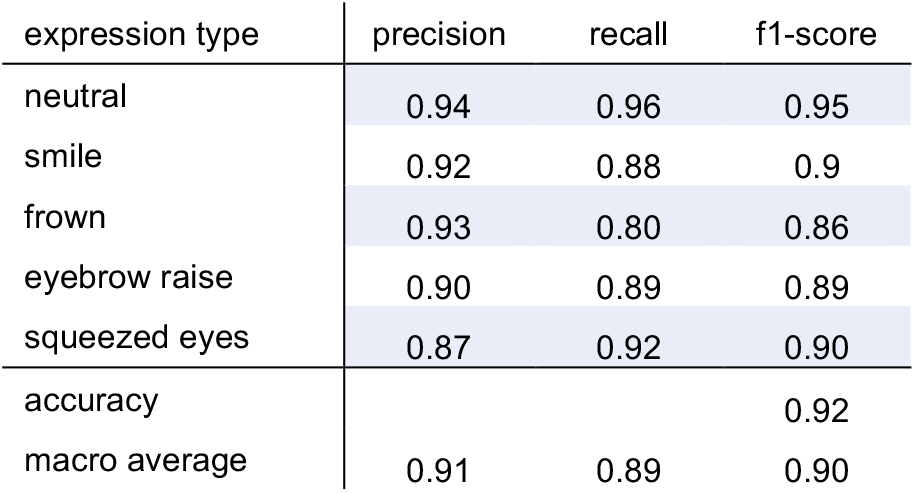
Classification report for the ML facial expression recognition model evaluated on the facial expressions performed with head movement.

**Figure 10.**
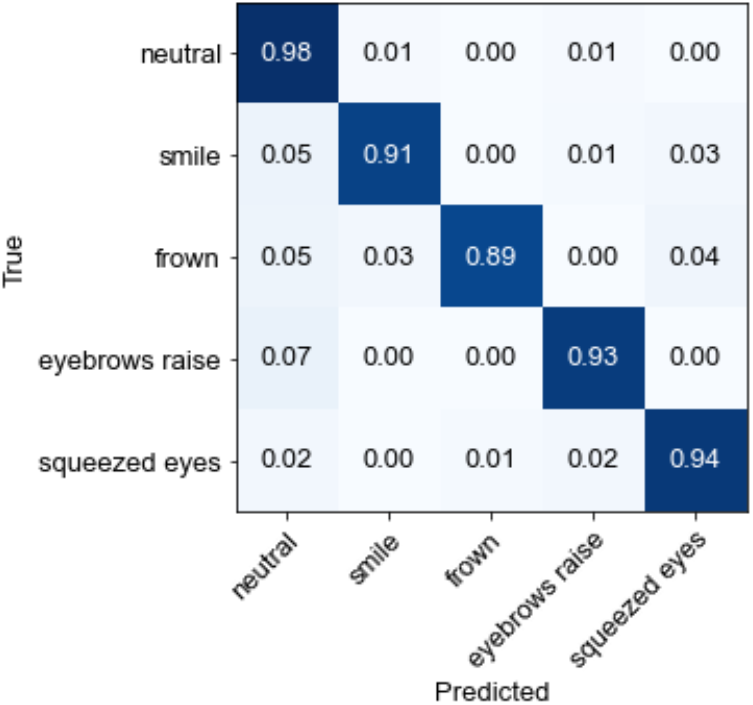
Confusion matrix for the ML facial expression recognition model evaluated on the facial expressions performed in a still scenario.

**Figure 11.**
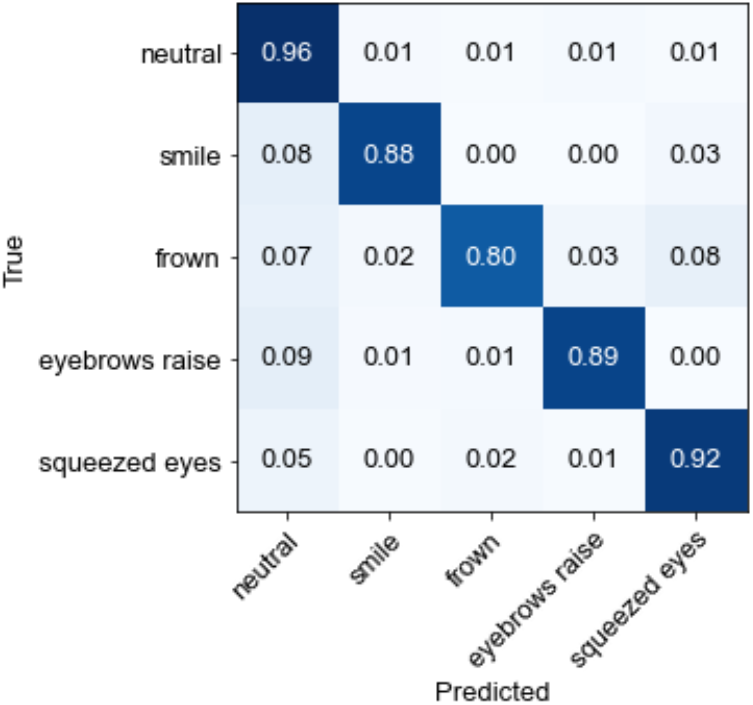
Confusion matrix for the ML facial expression recognition model evaluated on the facial expressions performed with head movement.

#### Low vs. medium vs. high position of the glasses

To evaluate the influence of the positioning of the glasses on the model’s performance in recognizing facial expressions, separate analyses were conducted on expressions performed while the participants were wearing the glasses in three different positions: low (the lowest point on the nasal bridge where the glasses are staying in place), medium (the medium point on the nasal bridge), and high (the highest point on the nasal bridge). These comparisons include only high-intensity expressions with varying duration (short and long). The classification matrices for the low, medium, and high position are illustrated in Figure 12, Figure 13, and Figure 14, respectively. The classification reports are also presented in Table 6, Table 7, and Table 8. The model demonstrates high accuracy in recognizing the facial expressions, regardless of the position of the glasses. Namely, the model achieved an accuracy of 92% on the low-position and medium-position data (f1-score of 0.88 and 0.87, respectively), and an accuracy of 94% on the high-position data (f1-score of 0.9). The increased accuracy observed in the high-position scenario in comparison to the low and medium position can be attributed to the reduction in the number of instances of facial expressions being misclassified as neutral. This effect is particularly prominent for the frown and eyebrow raise expression. This can be explained by the possibility that the sensors located on the upper frame of the glasses to not be able to capture skin movement and muscle activation in the eyebrow and forehead area, which are the regions primarily involved in these two expressions, when the glasses are worn in the low or medium position. The model’s performance for the recognition of other expressions remains consistent across all positions of the glasses.

**Table 6.**
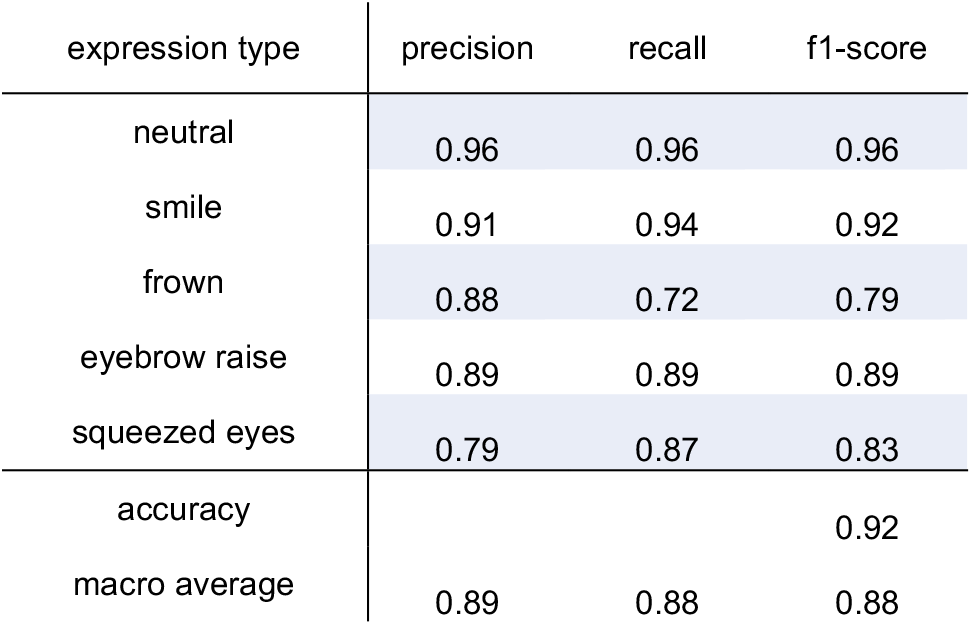
Classification report for the ML facial expression recognition model evaluated on the ‘low-position’ data.

**Table 7.**
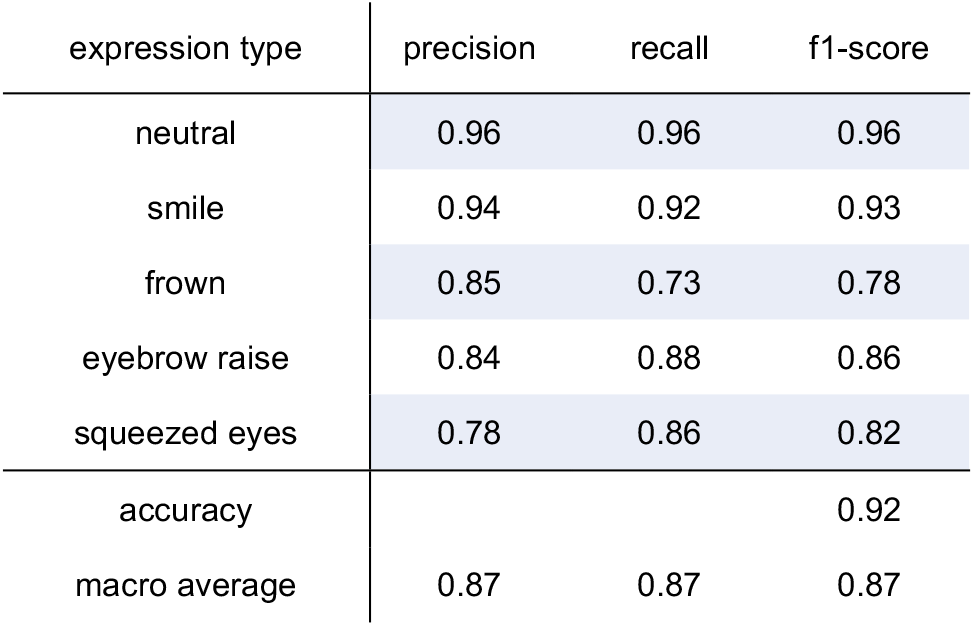
Classification report for the ML facial expression recognition model evaluated on the ‘medium-position’ data.

**Table 8.**
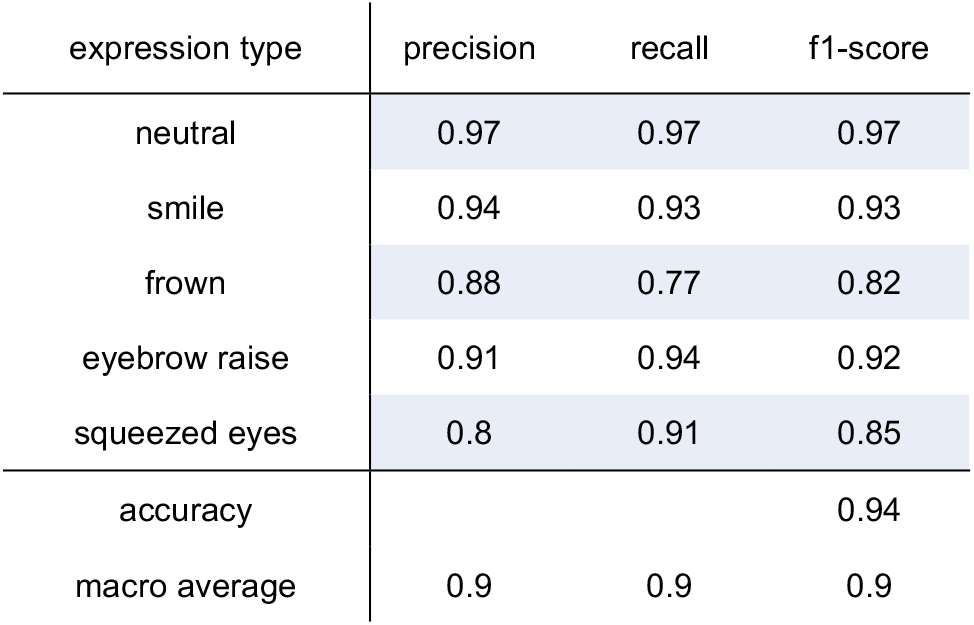
Classification report for the ML facial expression recognition model evaluated on the ‘high-position’ data.

**Figure 12.**
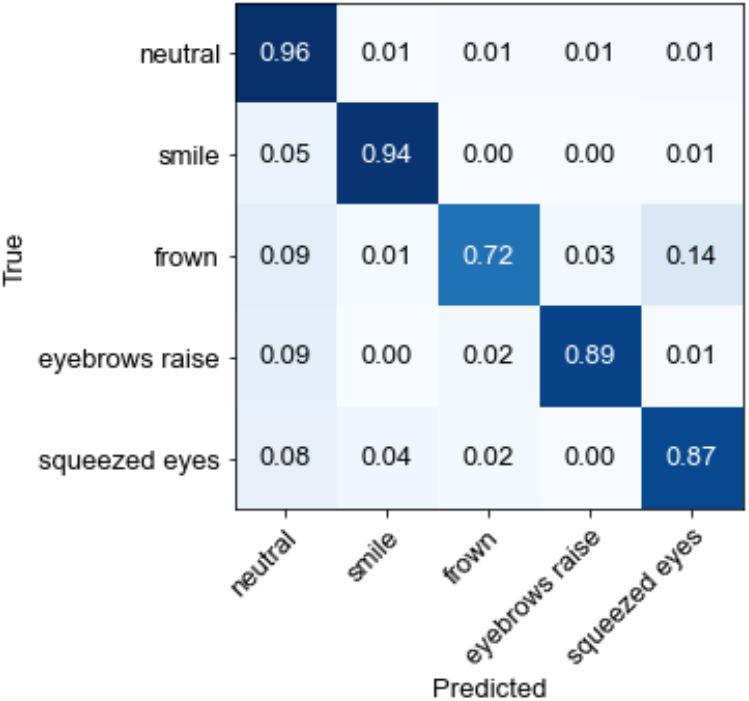
Confusion matrix for the ML facial expression recognition model evaluated on the ‘low-position’ data.

**Figure 13.**
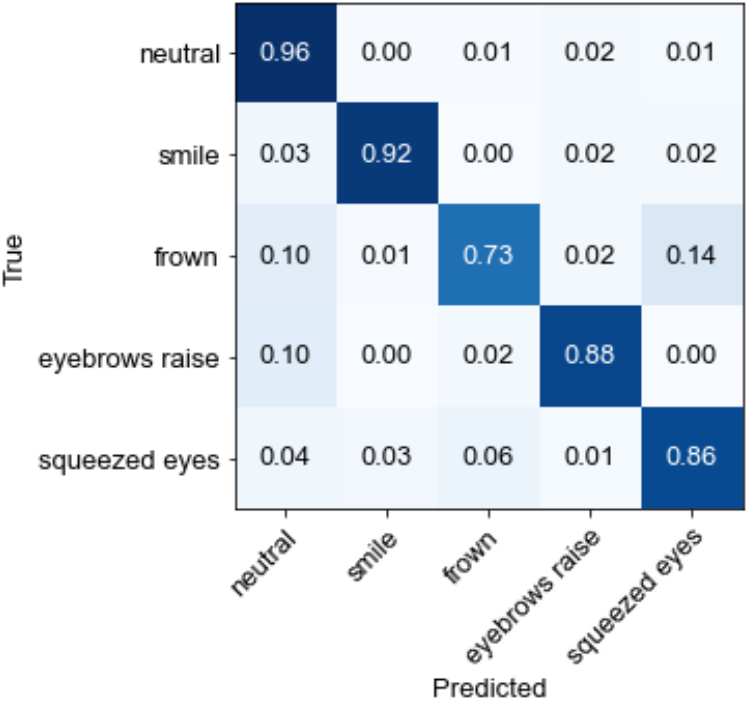
Confusion matrix for the ML facial expression recognition model evaluated on the ‘medium-position’ data.

**Figure 14.**
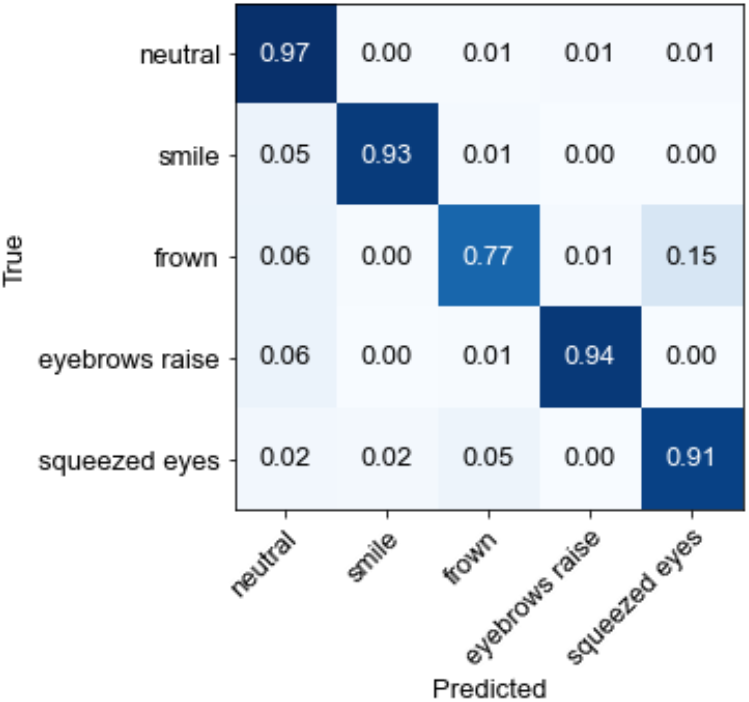
Confusion matrix for the ML facial expression recognition model evaluated on the ‘high-position’ data.

## Discussion

In this study we explored the performance of OMG-based smart glasses (OCOsense™) for monitoring facial movements associated with different facial expressions. We performed statistical analysis and machine learning analysis.

The results from the statistical analysis indicated that:

- OCO™ sensors were able to accurately capture differences in cheek and brow movements with a high level of sensitivity and specificity. This includes detecting both low-intensity expressions and high-intensity expressions (Figure 2 and Figure 3). This is particularly noteworthy, as previous studies have shown that subtle expressions made at a low intensity can be challenging to recognize and differentiate, even for the human eye [25, 26, 27]. This highlights that OCOsense™ may be a valuable tool for monitoring both intense and subtle facial expressions, which convey potentially valuable information about an individual’s emotional state.
- The analysis on the influence of head movement on the OCO™ sensor data showed that head movements did not have a statistically significant impact on the sensor data in the explored scenario. This is an important finding, as it suggests that OCOsense glasses may be well-suited for monitoring facial expressions in real-world settings, where head movement obviously will be expected to co-occur with facial expressions. Nevertheless, additional mechanisms based on the sensor data can be employed in order to avoid faulty sensor reading. For example, one could avoid sensor readings above 30 mm as this is outside of known specification for the OCO™ sensors.
- The position of the glasses was found to have some influence on the OCO™ sensors measurements, particularly when monitoring the brow movement during frown, eyebrow raise, and squeezed eyes expression. This can be explained by the possibility that the sensors located on the upper frame of the glasses are unable to capture skin movement in the eyebrow and forehead area, which are the regions primary involved in these three expressions, when the glasses are not correctly positioned. This finding highlights the importance proper fit of the glasses on the wearer’s face to ensure accurate measurements are taken.

The results from the statistical analysis were further confirmed with the machine learning experiments. The measurements obtained from the OCO™ sensors provided valuable data for the development of a robust machine learning model that can differentiate between a neutral state and four different facial expressions performed in different conditions, including variations in the intensity, duration, and presence of head movement, as well as glasses position. Depending on the conditions, the f1-score varied between .87 and .93.

### Conclusions and implications for future work

In conclusion, this study evaluated the performance of the OCOsense™ smart glasses with novel OMG based OCO™ sensors, which measure skin movement in three dimensions over key facial muscles, as a tool for capturing facial expressions and recognition of those expressions using statistical and machine learning methods. The experiments were conducted using a dataset gathered from 27 participants, who were performing facial expressions varying in intensity, duration, and head movement. Statistical tests showed expected differences in the cheek and the brow movement during smile, frown, eyebrow raise, and squeezed eyes expression – namely, the sensors detected increased cheek skin movements during smile and squeezed eyes expression compared to frown and eyebrow raise expression. Conversely, increased brow movement was detected during the eyebrow raise and frown expression, compared to the squeezed eyes and smile expressions. The study also found that head movements do not have a significant impact on the measurements obtained from the OCO™ sensors when monitoring the movement of the cheek and brow during different facial expressions. However, the position of the glasses was found to have some influence on the OCO™ sensors measurements, particularly when monitoring the movement of the brow during frown, eyebrow raise, and squeezed eyes expressions. Furthermore, the use of the OCO™ sensors provided valuable data for the development of a ML-based expression recognition algorithm, which yielded an accuracy of 93% (0.90 f1-score) evaluated on data collected from n=27 participants using a leave-one-subject-out cross-validation technique.

Collectively, these findings demonstrate that the expression recognition model based on the OCOsense™ glasses data can be used as a reliable tool for monitoring facial expressions as well as serving to highlight the potential of this technology in a variety of applications. For example, in the field of human-computer interaction, the use of smart glasses with an expression recognition model can enable more natural and intuitive interactions between individuals and technology. Recognized expressions can be used as a source for interaction and control of different devices, such as hands-free control of a wheelchair or hands-free control of a heads-up display, without the need to interact with physical buttons. As facial expressions play a major role in conveying affective states, such sensing technology can be used in the emotion recognition and affective computing field within real-world scenarios or longitudinal studies for continuous monitoring and managements of emotion disorders. For mental health professionals, this technology could provide an objective way to monitor facial expressivity, as one of the major behavioural markers of depression [28]. This can aid them in gaining deeper insights and understanding of their patients’ emotional states, enabling them to provide more personalized treatment that addresses specific needs.

The next steps in this research include moving from analysing voluntary facial expressions to analysing spontaneous expressions and robustly validating the glasses’ ability to detect them in naturalistic environments. This will involve testing the glasses on a diverse group of participants to ensure that the detection of expressions is not affected by factors such as race, gender, or age. Furthermore, the research will also involve further development of the model to improve its ability to detect subtle spontaneous expressions and gestures, which are often harder to detect but may still convey valuable information about a person’s emotional state.

## Methods

### Participants

A group of 27 healthy volunteers, 11 females and 14 males, with a mean age of 26.3 ± 7.66 (range 16 – 47) were recruited to participate in the experiment. The inclusion age range was 16 – 68 years. Exclusion criteria for recruitment were the presence of facial neuromuscular and nervous disorders. Ethical approval was obtained from the London - Riverside Research Ethics Committee on 15 July 2022 (ref: 22/LIO/0415). After a detailed explanation of the experimental procedure, all participants also provided written informed consent before participating in the study. The experiment was conducted following institutional ethical provisions and the Declaration of Helsinki.

### Apparatus

For data collection, we used the Emteq’s OCOsense™ smart glasses. The glasses contain six OCO™ sensors, a 9-axis IMU, altimeter and integrated dual speech detection microphones. In this study, we analysed only the data coming from the OCO™ sensors. The OCO™ sensors are proprietary sensors developed by Emteq Labs, which use an optical non-contact approach called optomyography (OMG). They are used to measure skin movement due to underlying myogenic activity in 3 dimensions (X, Y, and Z) and do not require skin contact, i.e., they can function accurately from 4 mm to 30 mm away from the skin. The sensors are built into the OCOsense™ glasses frame and are positioned to measure skin movement over the frontalis muscle (left and right side of the forehead), zygomaticus major muscle (left and right side of the cheeks), and the left and right temples.

### Experimental scenario

Prior to initiation of the data collection procedure, all participants were briefed on the experimental design, task requirements, and equipment utilized.

The data collection protocol involved performing three different tasks (Task A, Task B, and Task C) in which the participants were instructed to perform four distinct facial expressions, namely, smile, frown, eyebrow raise, and squeezed eyes, varying in intensity and duration. All expressions were repeated three times in each task:

1. In task A, the participants were required to perform voluntary expressions with varying intensity (low and high) and duration (short and long) as follows: 1) low-intensity, short-duration expressions (one second per repetition); 2) low-intensity, long duration expressions (three seconds per repetition); 3) high-intensity, short duration expressions (one second per repetition); and 4) high-intensity, long duration expressions (three seconds per repetition).
2. In Task B, the participants were required to perform voluntary expressions of high intensity and long duration while simultaneously moving their head in a specific direction, as follows: 1) high-intensity expressions with head movement to the left (three seconds per repetition); 2) high-intensity expressions with head movement to the right (three seconds per repetition); 3) high-intensity expressions with head movement upwards (three seconds per repetition); and 4) high--intensity expressions with head movement downwards (three seconds per repetition).
3. In task C, the participants were required to perform voluntary expressions of high-intensity and short and long duration while wearing the glasses in three different positions: low (the lowest point on the nasal bridge where the glasses are staying in place), medium (the medium point on the nasal bridge), and high (the highest point on the nasal bridge). The expressions in Task C were performed in the following order: 1) low position: high-intensity, short duration expressions (one second per repetition); 2) low position: high-intensity, long duration expressions (three seconds per repetition); 3) medium position: high-intensity, short duration expressions (one second per repetition); 4) medium position: high-intensity, long duration expressions (three seconds per repetition); 5) high position: high-intensity, short duration expressions (one second per repetition); and 6) high position: high-intensity, long duration (three seconds per repetition).

The data collection process was uninterrupted. The participants were instructed to maintain a neutral facial expression in between each posed expression.

### Statistical analysis

#### Data preprocessing

The following data processing steps were applied to the acquired sensor data:

- Linear trend removal: The linear trend was removed from each signal obtained from the OCO™ cheek and brow sensors to eliminate the influence of long-term drift on the measured skin movement.
- Calculation of the vector magnitude for each sensor: As the OCO™ sensors measure skin movement in 3 dimensions (X, Y, and Z), the vector magnitude was calculated for each sensor 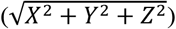.
- Combination of processed sensor signals values from the left and right sensors: The vector magnitude value from the left cheek sensor was added to the vector magnitude value from the right cheek sensor, and the same was done for the brow sensors. This resulted in the creation of two signals, one representing the total cheek movement (left + right), and one representing the total brow movement (left + right).
- Smoothing of the resulting signals: The resulting cheek and brow signals were smoothed using a rolling median filter with a window size of 15 samples to reduce the effects of noise on the signals.

#### Statistical tests

The overall data analysis was conducted using Python programming language. Hypothesis testing was performed using the Wilcoxon signed-rank test. The Wilcoxon signed-rank test is a non-parametric version of the paired T-test that compares the distribution of differences between related paired samples to determine if they come from the same distribution. The null hypothesis of the test is that the samples come from the same distribution. The *p*-values were Bonferroni-corrected. The significance annotations were represented as: * if *p* ∈ [.05, 10^−2^); ** if *p* ∈ [10^−2^, 10^−3^); *** if *p* ∈ [10^−3^, 10^−4^); and **** *if p* >=10^−4^.

### OCOsense™ Expression Recognition Machine Learning Pipeline

To model the relation between the facial movements detected by the OCO™ sensors and the facial expressions, we used a simple expression recognition algorithm utilizing signal processing and machine learning techniques. The algorithm utilized data from all six OCO™ sensors incorporated in the OCOsense™ glasses, resulting in a total of 18 sensor streams. During the data collection procedure, the OCO™ data were continuously recorded at a fixed rate of 50 Hz. The raw sensor data underwent a data preprocessing pipeline, including filtering, segmentation, and feature engineering. The data were filtered using a low-pass filter, and the linear trend was removed from each signal stream. A sliding window technique was utilized for the sensor data segmentation. The signals were segmented using a 0.1-second window and a 0.1-second window stride, meaning that there was no overlap between consecutive windows. Eventually, 9 statistical features per sensor stream were extracted, resulting in a total of 162 features. The features included the mean, standard deviation, minimum, maximum, range, interquartile range, kurtosis, skewness, and root mean square. The feature set for each participant was also standardized (removing the mean and scaling to unit variance). The standardized features were used as an input to a Random Forest algorithm, which output the recognized expression as smile, frown, eyebrow raise, squeezed eyes, or neutral state. Due to the imbalanced nature of the dataset, the neutral class was under sampled in the training process, so that all five classes were evenly distributed.

## Data Availability

The data is available on demand.

https://www.emteqlabs.com/

## Acknowledgements

The authors acknowledge the contributions of all their colleagues at Emteq Ltd. towards the development of the OCOsense™ system. Special thanks go to Bojan Jakimovski, Borjan Sazdov, and Bojan Sofronievski for their support in the data collection and data preparation process for the study.

## Data Availability

The datasets generated and analysed during the current study are not publicly available due to company policies on data protection and confidentiality.

## Author contribution

All authors have approved of the submitted version of this manuscript and have agreed to be accountable for their contributions and for the work in this manuscript. The authors specific contributions were as follows: Writing (review and editing), conceptualization and interpretation: all authors; Formal analysis: I.K; Investigation, methodology, visualization, and software: I. K., M. G., S. S., J.A., M.F.; Project administration, Funding acquisition: M. J. B., H. G., and C. N.

## Competing interests statement

Dr. M. Gjoreski and dr. H. Gjoreski have previously worked as consultants with the funding company, though they did not receive funding for this study. I. Kiprijanovska, S. Stankoski, J. Archer, M. Fatoorechi, and dr. M. J. Broulidakis are employees at the funding company. Dr. C. Nduka MA, MD, FRCS, is the founder of the funding company.

## Funding

This work was supported by Innovate UK under the project “Mobile Observation of Depression (MOOD) platform for digital phenotyping” (Grant number 105207). H. Gjoreski’s work was partially funded by the WideHealth project (EU’s Horizon 2020 research and innovation programme, grant agreement No 952279).

## Notes

### Author Declarations

Ethical approval was obtained from the London - Riverside Research Ethics Committee on 15 July 2022 (ref: 22/LIO/0415).

